# Systematic Review of the Registered Clinical Trials of Coronavirus Disease 2019 (COVID-19)

**DOI:** 10.1101/2020.03.01.20029611

**Authors:** Rui-fang Zhu, Yu-lu Gao, Sue-Ho Robert, Jin-ping Gao, Shi-gui Yang, Chang-tai Zhu

## Abstract

**Background:** Since the outbreak of coronavirus disease 2019 (COVID-19), many researchers in China have immediately carried out clinical research scheme of the COVID-19. But, there is still a lack of systematic review of registered clinical trials. Therefore, we conducted a systematic review of the clinical trials of COVID-19 to summarize the characteristics of the COVID-19 registered clinical trials.

**Methods:** This study is based on the recommendations of the PRISMA in the Cochrane handbook. The databases from the Chinese Clinical Registration Center and the ClinicalTrials.gov were searched to collect the registered clinical trials of COVID-19. The retrieval inception date is February 9, 2020. Two researchers independently selected the literature based on inclusion and exclusion criteria, extracted data and evaluated the risk of bias.

**Results:** A total of 75 registered clinical trials (63 interventional studies and 12 observational studies) of COVID-19 were obtained. A majority of clinical trials were sponsored by Chinese hospitals. Only 11 trials have begun to recruit patients, and none of the registered clinical trials had been completed; 34 trials were early clinical exploratory trials or in a pre-experiment stage, 15 trials belonged to phrase III and 4 trials were phrase IV. The methods of intervention included traditional Chinese medicine involving 26 trials, Western medicine involving 30 trials, and integrated traditional Chinese medicine and Western medicine involving 19 trials. The subjects were mainly non-critical adult patients (≥ 18 years old). The median sample size of the trials was 100 (IQR: 60 - 200), and the median execute time of the trials was 179 d (IQR: 94 - 366 d). The main outcomes were clinical observation and examinations. Overall, both the methodology quality of interventional trials and observational studies were low.

**Conclusions:** Disorderly and intensive clinical trials of COVID-19 using traditional Chinese medicine and western medicine are ongoing or will being carried out in China. However, based on the low methodology quality and small sample size and long studies execute time, we will not be able to obtain reliable, high-quality clinical evidence about COVID-19 treatment in the near future. Improving the quality of study design, prioritizing promising drugs, and using different designs and statistical methods are worth advocating and recommending for the clinical trials of COVID-19 in China.

---

Coronavirus disease 2019 (COVID-19), being an emerging infectious disease, is a serious threat to human health [1-3]. In December 2019, the initial outbreak of COVID-19 in Wuhan city, Hubei province of China, was suspected to be related to the seafood market, and chrysanthemum head bat was suspected to be the host of the new coronavirus [4-7]. Patients with COVID-19 show manifestations of respiratory tract infection, such as fever, cough, pneumonia, and in severe cases, death [8, 9]. According to a recent survey, the mortality rate of the viral disease is estimated to be about 2% - 4% [8, 10]. By Feb 29, 2020, more than 80,000 people were confirmed to be infected around the world, with most of them belonging to China. At present, there are different number of infected people in different provinces of China, with Hubei Province being the most seriously affected one), and the signs of infection outbreak are obvious. In addition, more than 40 countries around the world have also seen new cases of COVID-19 [11-15]. Therefore, COVID-19 is a great challenge to human health [10, 16].

Little is known about COVID-19 as it is a new infectious disease; therefore, presently there is no specific treatment available for COVID-19. To date, no clinical intervention trial has been completed and reported. Due to the urgent need for treatment prevention and control of the disease, it is necessary to develop effective intervention methods for COVID-19 to facilitate disease control. Since the outbreak of the COVID-19, many researchers in China have carried out clinical research trials, aiming to develop strategies for the treatment, prevention and diagnosis of COVID-19. However, up to now, there is still a lack of systematic appraisal of the registered clinical trials COVID-19. Therefore, we conducted a systematic review of the clinical trials of COVID-19 to analyze the characteristics and existing problems of the registered clinical trials.

## Materials and methods

### Inclusion criteria

This review was performed according to the Cochrane Handbook for Systematic Reviews of Interventions [17] and presented based on Preferred Reporting Items for Systematic Reviews and Meta-analyses guidelines [18].

The inclusion criteria of this study were: patients with COVID-19; clinical trials with protocol; trials involving the diagnosis, prevention and treatment of COVID-19; trials having clear and specific end-point outcomes; and trials with any type of study design.

### Exclusion criteria

The exclusion criteria of this study were: animal trials; theoretical research; and unregistered clinical trials.

### Retrieval strategies

The literature retrieval was independently completed by two researchers. The databases from the Chinese clinical trial registration center and the ClinicalTrials.gov were used for data search. No language limitations were specified for the search, and the search deadline was February 9, 2020. The following key words were applied: new coronavirus, COVID-19, 2019-nCoV pneumonia, novel coronavirus pneumonia, 2019-nCoV infection, new coronavirus infection, new coronavirus, etc.

### Data extraction

The contents that were extracted mainly included registration number, project name, research leader, research type, study design, sponsor, implementation unit, start time, completion period, research site, research institute, stage, research object, inclusion standard, exclusion standard, sample size, setting, location, recruitment period, intervention group measures, control group measures, random methods, blind methods, distribution concealment and measurement indicators. Literature evaluation was independently conducted by two researchers.

### Methodology quality assessment

The quality evaluation and data extraction of each literature fulfilling the inclusion criteria was conducted independently and a cross-check was carried out. Arguments or disagreement of opinions were resolved by a discussion between the two researchers. The randomized controlled trial was based on Cochrane risk of bias items, which includes: randomization sequence generation, allocation concealment, blinding of participants and personnel, blinding of outcome assessment, incomplete outcome data, selective reporting, and other bias [19]. The observational study was based on the quality evaluation by Newcastle-Ottawa scale (NOS) [20].

### Summary and synthesis

This review presents a narrative synthesis. This study mainly analyzes and summarizes the types of studies, intervention, host organization and address, sample size, research stage, research status, expected completion time, inclusion and exclusion criteria, outcome measurement and observation time, and methodology quality and describes the results with statistics and characteristics. Non-parametric data are represented by median and 95% CI and the statistical analysis used MedCalc statistical software (version 15.2.2, MedCalc Software bvba, Ostend, Belgium; http://www.medcalc.org; 2015). The bias plot was performed by Review Manager (RevMan) [Computer program] (version 5.2, Copenhagen: The Nordic Cochrane Centre, The Cochrane Collaboration, 2012).

## Results

### Trial search results

Up to February 9, 2020, we retrieved a total of 75 clinical trials of COVID-19 from the Chinese clinical registration center, and 18 clinical trials of COVID-19 from the ClinicalTrails.gov, and a total of 75 clinical trials of COVID-19 were obtained (Table 1 and Table 2). The retrieval process is shown in Figure 1.

**Table 1.** Summary of intervention registered clinical trials.

| N<br>o | Register number | Study<br>leader<br>(year) | Primary sponsor | Study name |
| --- | --- | --- | --- | --- |
| 1 | ChiCTR2000029638 | Liu L<br>2020 | West China Hospital,<br>Sichuan University | Multicenter randomized controlled trial for novel recombinant high-efficiency compound interferon in the treatment of novel coronavirus pneumonia (COVID-19) |
| 2 | ChiCTR2000029387 | Chen Y<br>2020a | Chongqing Public Health Medical Center | Comparison of efficacy and safety of three antiviral regimens in patients with mild to moderate novel coronavirus pneumonia (COVID-19): a randomized controlled trial |
| 3 | ChiCTR2000029386 | Chen Y<br>2020b | Chongqing Public Health Medical Center | Adjunctive corticosteroid therapy for Patients with Severe Novel coronavirus pneumonia (COVID-19): a randomized controlled trial |
| 4 | ChiCTR2000029435 | Wei L<br>2020 | Wuhan First Hospital | Randomized controlled trial for traditional Chinese medicine in the prevention of novel coronavirus pneumonia (COVID-19) in high risk population |
| 5 | ChiCTR2000029308 | Huang C<br>2020 | Wuhan Jinyintan Hospital (Wuhan Infectious Diseases Hospital) | A randomized, open-label, blank-controlled trial for the efficacy and safety of lopinavir-ritonavir and interferon-alpha 2b in hospitalization patients with novel coronavirus pneumonia (COVID-19) |
| 6 | ChiCTR2000029400 | Hung L<br>2020 | China Academy of Chinese Medical Sciences | Clinical controlled trial for traditional Chinese medicine in the treatment of novel coronavirus pneumonia (COVID-19) |
| 7 | ChiCTR2000029418 | Liang T<br>2020 | Dongzhimen Hospital<br>Affiliated to Beijing University of Chinese Medicine | Chinese herbal medicine for severe novel coronavirus pneumonia (COVID-19): a randomized controlled trial |
| 8 | NCT04244591 | Du B<br>2020 | Peking Union Medical College Hospital | Glucocorticoid therapy for novel coronavirus critically ill patients with severe acute respiratory failure |
| 9 | NCT04251871 | Wang R<br>2020 | Beijing 302 Hospital | Treatment and prevention of traditional Chinese medicines (TCMs) on 2019-nCoV infection |
| 10 | ChiCTR2000029436 | Li J 2020 | The First Hospital of He'nan University of Chinese Medicine | A single arm study for evaluation of integrated traditional Chinese and western medicine in the treatment of novel coronavirus pneumonia (COVID-19) |
| 11 | ChiCTR2000029432 | Yang Z<br>2020 | The First Affiliated Hospital of Guangzhou University of Chinese Medicine | A real world study for the efficacy and Safety of large dose tanreqing injection in the treatment of patients with novel coronavirus pneumonia (COVID-19) |
| 12 | ChiCTR2000029431 | Zhao D<br>2020 | Affiliated Zhongshan Hospital of Dalian University | Clinical study for the remedy of M1 macrophages target in the treatment of novel coronavirus pneumonia (COVID-19) |
| 13 | ChiCTR2000029381 | Zhong N<br>2020 | The First Affiliated Hospital of Guangzhou Medical University | A prospective comparative study for Xue-Bi-Jing injection in the treatment of novel coronavirus pneumonia (COVID-19) |
| 14 | ChiCTR2000029487 | Su W<br>2020 | Wuhan Hospital of Integrated Traditional Chinese and | Clinical study for Gu-Biao Jie-Du-Ling in preventing of 2019-nCoV pneumonia (Novel coronavirus pneumonia, NCP) |
|  |  |  | Western Medicine | in children |
| 15 | ChiCTR2000029479 | Tang J<br>2020 | Hospital of Chengdu University of Traditional Chinese Medicine | Research for traditional Chinese medicine technology prevention and control of 2019-nCoV pneumonia (novel coronavirus pneumonia, NCP) in the community population |
| 16 | ChiCTR2000029468 | Jiang H<br>2020 | Sichuan Academy of Medical Sciences & Sichuan Provincial People's Hospital | A real-world study for lopinavir/ritonavir (LPV/r) and emtricitabine (FTC) / Tenofovir alafenamide Fumarate tablets (TAF) regimen in the treatment of 2019-nCoV pneumonia (novel coronavirus pneumonia, NCP) |
| 17 | ChiCTR2000029461 | Xia W<br>2020 | Xinhua affiliated hospital, Hubei University of Chinese Medicine | A Randomized Controlled Trial for Integrated Traditional Chinese Medicine and Western Medicine in the Treatment of Common Type 2019-nCoV Pneumonia (Novel Coronavirus Pneumonia, NCP) |
| 18 | ChiCTR2000029460 | Zheng C<br>2020 | Xinhua affiliated hospital, Hubei University of Chinese Medicine | The effect of shadowboxing for pulmonary function and quality of life in patients with 2019-nCoV pneumonia (novel coronavirus pneumonia, NCP) in rehabilitation period |
| 19 | ChiCTR2000029459 | Xia W<br>2020 | Xinhua affiliated hospital, Hubei University of Chinese Medicine | The effect of pulmonary rehabilitation for pulmonary function and quality of life in patients with 2019-nCoV pneumonia (novel coronavirus pneumonia, NCP) in rehabilitation period |
| 20 | ChiCTR2000029439 | Wang Y<br>2020 | Beijing hospital of Traditional Chinese medicine | Combination of traditional chinesemedicne and western medicine in the treatment of common type 2019-nCoV pneumonia (novel coronavirus pneumonia, NCP) |
| 21 | ChiCTR2000029438 | Liu Q 2020 | Hubei integrated traditional Chinese and Western Medicine Hospital | A randomized controlled trial of integrated TCM and Western Medicine in the treatment of severe 2019-nCoV pneumonia (novel coronavirus pneumonia, NCP) |
| 22 | NCT04252274 | Lu 2020a | Shanghai Public Health Clinical Center | Efficacy and Safety of Darunavir and Cobicistat for Treatment of Pneumonia Caused by 2019-nCoV |
| 23 | NCT04261517 | Lu 2020b | Shanghai Public Health Clinical Center | Efficacy and Safety of Hydroxychloroquine for Treatment of Pneumonia Caused by 2019-nCoV ( HC-nCoV ) |
| 24 | ChiCTR2000029544 | Qiu Y 2020a | The First Affiliated Hospital, Zhejiang University School of Medicine | A randomized controlled trial for the efficacy and safety of BaloxavirMarboxil, Favipiravir tablets in 2019-nCoV pneumonia (novel coronavirus pneumonia, NCP) patients who are still positive on virus detection under the current antiviral therapy |
| 25 | ChiCTR2000029542 | Jiang S 2020 | Sun Yat-sen Memorial Hospital, Sun Yat-sen University | Study for the efficacy of chloroquine in patients with 2019-nCoV pneumonia (novel coronavirus pneumonia, NCP) |
| 26 | ChiCTR2000029541 | Wang H 2020 | Zhongnan Hospital of Wuhan University | A randomised, open, controlled trial for darunavir/cobicistat or Lopinavir/ritonavir combined with thymosin a1 in the treatment of 2019-nCoV pneumonia (novel coronavirus pneumonia, NCP) |
| 27 | ChiCTR2000029539 | Zhao J 2020 | Tongji Hospital, Tongji Medical College, Huazhong University of Science and Technology | A randomized, open-label study to evaluate the efficacy and safety of Lopinavir-Ritonavir in patients with mild 2019-nCoV pneumonia (novel coronavirus pneumonia, NCP) |
| 28 | ChiCTR2000029518 | Wen C<br>2020a | Zhejiang Chinese Medical University | Chinese medicine prevention and treatment program for 2019-nCoV pneumonia (novel coronavirus pneumonia, NCP): a perspective, double-blind, placebo, randomised controlled trial |
| 29 | ChiCTR2000029517 | Wen C<br>2020b | Zhejiang Chinese Medical University | Chinese medicine prevention and treatment program for suspected 2019-nCoV pneumonia (novel coronavirus pneumonia, NCP): a perspective, double-blind, placebo, randomised controlled trial |
| 30 | ChiCTR2000029496 | Gong G<br>2020 | The Second Xiangya Hospital of Central South University | A randomized, open label, parallel controlled trial for evaluating the efficacy of recombinant cytokine gene-derived protein injection in eliminating novel coronavirus in patients with 2019-nCoV pneumonia (novel coronavirus pneumonia, NCP) |
| 31 | ChiCTR2000029495 | Huang M<br>2020 | Xinhua affiliated hospital, Hubei University of Chinese Medicine | Traditional Chinese Medicine, Psychological Intervention and Investigation of Mental Health for Patients With 2019-nCoV Pneumonia (Novel Coronavirus Pneumonia, NCP) in Convalescent Period |
| 32 | ChiCTR2000029493 | Zhang J<br>2020 | Xinhua affiliated hospital, Hubei University of Chinese Medicine | Traditional Chinese Medicine for Pulmonary Fibrosis, Pulmonary Function and Quality of Life in Patients With 2019-nCoV Pneumonia (Novel Coronavirus Pneumonia, NCP) in Convalescent Period: a Randomized Controlled Trial |
| 33 | ChiCTR2000029580 | Zhou J<br>2020 | Tongji Hospital, Tongji Medical College, Huazhong University of | A prospective, single-blind, randomized controlled trial for Ruxolitinib combined with mesenchymal stem cell infusion in the treatment of |
|  |  |  | Science and Technology | patients with severe 2019-nCoV pneumonia (novel coronavirus pneumonia, NCP) |
| 34 | ChiCTR2000029589 | Liu Q 2020b | Beijing Hospital of Traditional Chinese Medicine | An open, prospective, multicenter clinical study for the efficacy and safety of Reduning injection in the treatment of 2019-nCoV pneumonia (novel coronavirus pneumonia, NCP) |
| 35 | ChiCTR2000029600 | Liu Y 2020 | The Third People's Hospital of Shenzhen | Clinical study for safety and efficacy of Favipiravir in the treatment of 2019-nCoV pneumonia (novel coronavirus pneumonia, NCP) |
| 36 | ChiCTR2000029601 | Tong X 2020a | Hubei Provincial Hospital of TCM | Community based prevention and control for Chinese medicine in the treatment of 2019-nCoV pneumonia (novel coronavirus pneumonia, NCP) in the isolate suspected and confirmed population |
| 37 | ChiCTR2000029602 | Tong X 2020b | Hubei Provincial Hospital of TCM | Clinical study for community based prevention and control strategy of novel coronavirus pneumonia (COVID-19) in the isolate suspected and confirmed population |
| 38 | ChiCTR2000029603 | Qiu Y 2020a | The First Affiliated Hospital, Zhejiang University School of Medicine | A Randomized, Open-Label, Multi-Centre Clinical Trial Evaluating and Comparing the Safety and Efficiency of ASC09/Ritonavir and Lopinavir/Ritonavir for Confirmed Cases of 2019-nCoV Pneumonia (Novel Coronavirus Pneumonia, NCP) |
| 39 | ChiCTR2000029605 | Liu C 2020 | Tongji Hospital, Tongji Medical College, Huazhong University of Science and Technology | A randomized, open-label, blank-controlled, multicenter trial for Shuang-Huang-Lian oral solution in the treatment of 2019-nCoV pneumonia (novel coronavirus pneumonia, NCP) |
| 40 | ChiCTR2000029578 | Wen C 2020 | Zhejiang Chinese Medical University | Chinese medicine prevention and treatment program for 2019-nCoV pneumonia (novel coronavirus pneumonia, NCP): a perspective, sing-arm trial |
| 41 | ChiCTR2000029573 | Li L 2020 | Jiehua biotechnology (Qingdao) co. LTD | A multicenter, randomized, open-label, positive-controlled trial for the efficacy and safety of recombinant cytokine gene-derived protein injection combined with abidole, lopinavir/litonavir in the treatment of 2019-nCoV pneumonia (novel coronavirus pneumonia, NCP) patients |
| 42 | ChiCTR2000029572 | Pei B 2020a | Xiangyang First People's Hospital | Safety and efficacy of umbilical cord blood mononuclear cells in the treatment of severe and critically 2019-nCoV pneumonia (novel coronavirus pneumonia, NCP): a randomized controlled clinical trial |
| 43 | ChiCTR2000029569 | Pei B 2020b | Xiangyang First People's Hospital | Safety and efficacy of umbilical cord blood mononuclear cells conditioned medium in the treatment of severe and critically 2019-nCoV pneumonia (novel coronavirus pneumonia, NCP): a randomized controlled trial |
| 44 | ChiCTR2000029559 | Zhang Z2020 | Renmin Hospital of Wuhan University | Therapeutic effect of hydroxychloroquine on 2019-nCoV pneumonia (novel coronavirus pneumonia, NCP) |
| 45 | ChiCTR2000029558 | Xie C 2020a | Hospital of Chengdu University of Traditional Chinese Medicine | Recommendations of Integrated Traditional Chinese and Western Medicine for Diagnosis and Treatment of 2019-nCoV Pneumonia (Novel Coronavirus Pneumonia, NCP) in Sichuan Province |
| 46 | ChiCTR2000029550 | Xie C 2020b | Hospital of Chengdu University of Traditional Chinese Medicine | Recommendations for Diagnosis and Treatment of Influenza Patients in the Hospital of Chengdu University of Traditional Chinese Medicine Under the Raging of 2019-nCoV Pneumonia (Novel Coronavirus Pneumonia, NCP) |
| 47 | ChiCTR2000029549 | Xie C 2020c | Hospital of Chengdu University of Traditional Chinese Medicine | Recommendations of Integrated Traditional Chinese and Western Medicine for 2019-nCoV Pneumonia (Novel Coronavirus Pneumonia, NCP) |
| 48 | ChiCTR2000029548 | Qiu Y 2020b | The First Affiliated Hospital, Zhejiang University School of Medicine | Randomized, open-label, controlled trial for evaluating of the efficacy and safety of BaloxavirMarboxil, Favipiravir, and Lopinavir-Ritonavir in the treatment of 2019-nCoV pneumonia (novel coronavirus pneumonia, NCP) patients |
| 49 | NCT04260594 | QU 2020 | Ruijin Hospital | Clinical Study of Arbidol Hydrochloride Tablets in the Treatment of Pneumonia Caused by Novel Coronavirus |
| 50 | NCT04261907 | QIU 2020 | First Affiliated Hospital of Zhejiang University | Evaluating and Comparing the Safety and Efficiency of ASC09/Ritonavir and Lopinavir/Ritonavir for Novel Coronavirus pneumonia |
| 51 | NCT04257656 | Cao B 2020a | Capital Medical University | Severe 2019-nCoV Remdesivir RCT |
| 52 | ChiCTR2000029636 | Hu B 2020 | Union Hospital, Tongji Medical College, Huazhong University of Science and Technology | Efficacy and safety of aerosol inhalation of vMIP in the treatment of 2019-nCoV pneumonia (novel coronavirus pneumonia, NCP): a single arm clinical trial |
| 53 | ChiCTR2000029626 | Fang X<br>2020 | The First Affiliated Hospital, Zhejiang University School of Medicine | Immune Repertoire (TCR & BCR) Evaluation and Immunotherapy Research in Peripheral Blood of 2019-nCoV Pneumonia (Novel Coronavirus Pneumonia, NCP) Patients |
| 54 | ChiCTR2000029621 | Qu J<br>2020 | Ruijin Hospital, Shanghai Jiao Tong University School of Medicine | Clinical study of arbidol hydrochloride tablets in the treatment of 2019-nCoV pneumonia (novel coronavirus pneumonia, NCP) |
| 55 | ChiCTR2000029609 | Shan H<br>2020 | The Fifth Affiliated Hospital of Sun Yat-Sen University | A prospective, open-label, multiple-center study for the efficacy of chloroquine phosphate in patients with 2019-nCoV pneumonia (novel coronavirus pneumonia, NCP) |
| 56 | ChiCTR2000029606 | Li L<br>2020b | The First Affiliated Hospital, College of Medicine, Zhejiang University | Clinical Study for Human Menstrual Blood-Derived Stem Cells in the Treatment of Acute Novel Coronavirus Pneumonia (NCP) |
| 57 | ChiCTR2000029625 | Cai H<br>2020 | The First Affiliated Hospital, Zhejiang University School of Medicine | Construction of Early Warning and Prediction System for Patients with Severe / Critical 2019-nCoV Pneumonia (Novel Coronavirus Pneumonia, NCP) |
| 58 | NCT04263402 | Han M<br>2020a | Tongji Hospital | The Efficacy of Different Hormone Doses in 2019-nCoV Severe Pneumonia |
| 59 | NCT04254874 | Han M<br>2020 b | Tongji Hospital | A Prospective, Randomized Controlled Clinical Study of Interferon Atomization in the 2019-nCoV Pneumonia |
| 60 | NCT04261270 | Han M<br>2020 c | Tongji Hospital | A Randomized, Open, Controlled Clinical Study to Evaluate the Efficacy of ASC09F and Ritonavir for 2019-nCoV Pneumonia |
| 61 | NCT04252664 | Cao B<br>2020 b | Capital Medical<br>University | Mild/Moderate 2019-nCoV<br>Remdesivir RCT |
| 62 | NCT04261426 | Li 2020 | Peking Union<br>Medical College<br>Hospital | The Efficacy of Intravenous<br>Immunoglobulin Therapy for<br>Severe 2019-nCoV Infected<br>Pneumonia |
| 63 | NCT04255017 | Han M<br>2020d | Tongji Hospital | A Prospective,Randomized<br>Controlled Clinical Study of<br>Antiviral Therapy in the<br>2019-nCoV Pneumonia |

**Table 2.**
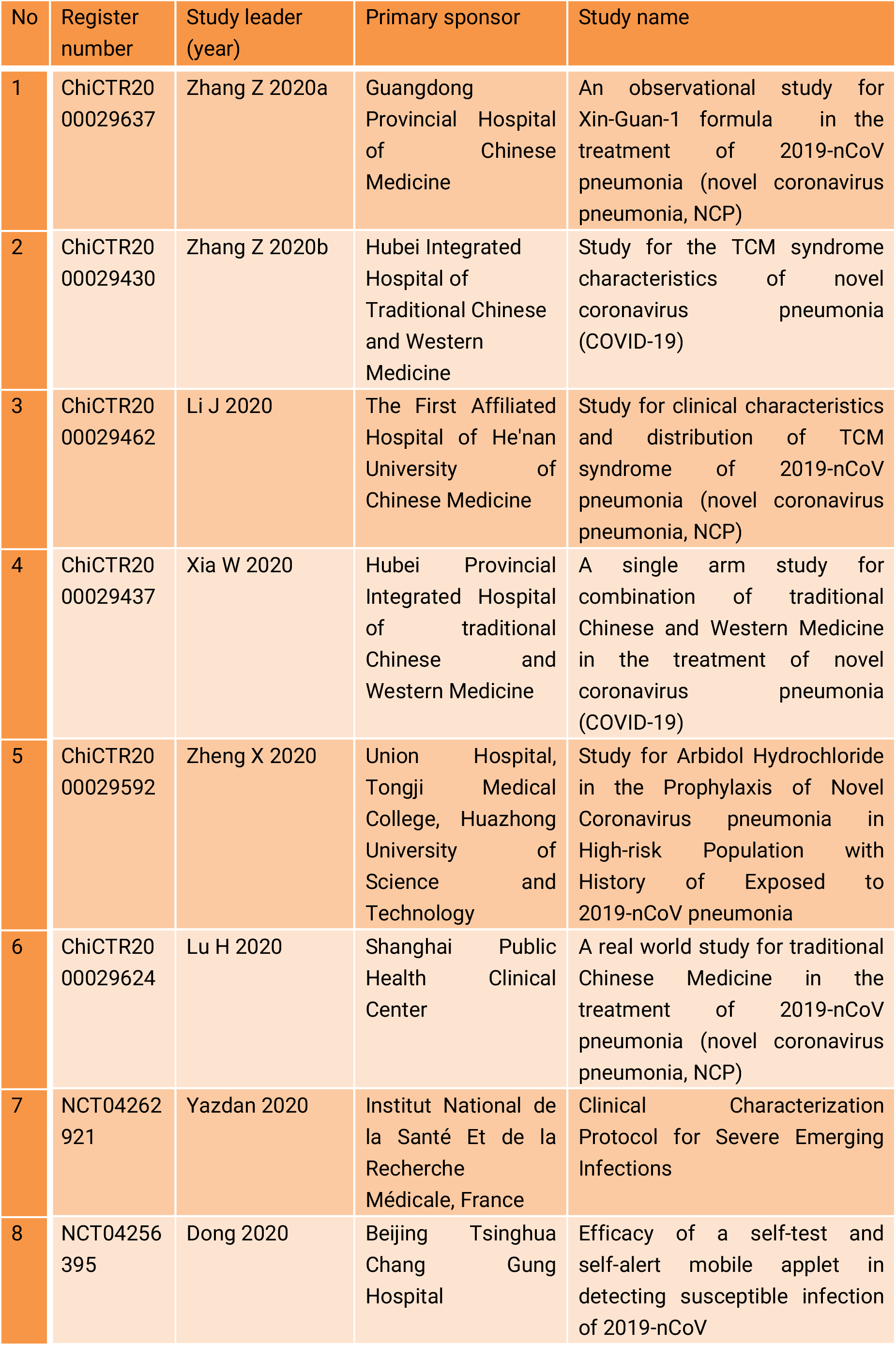
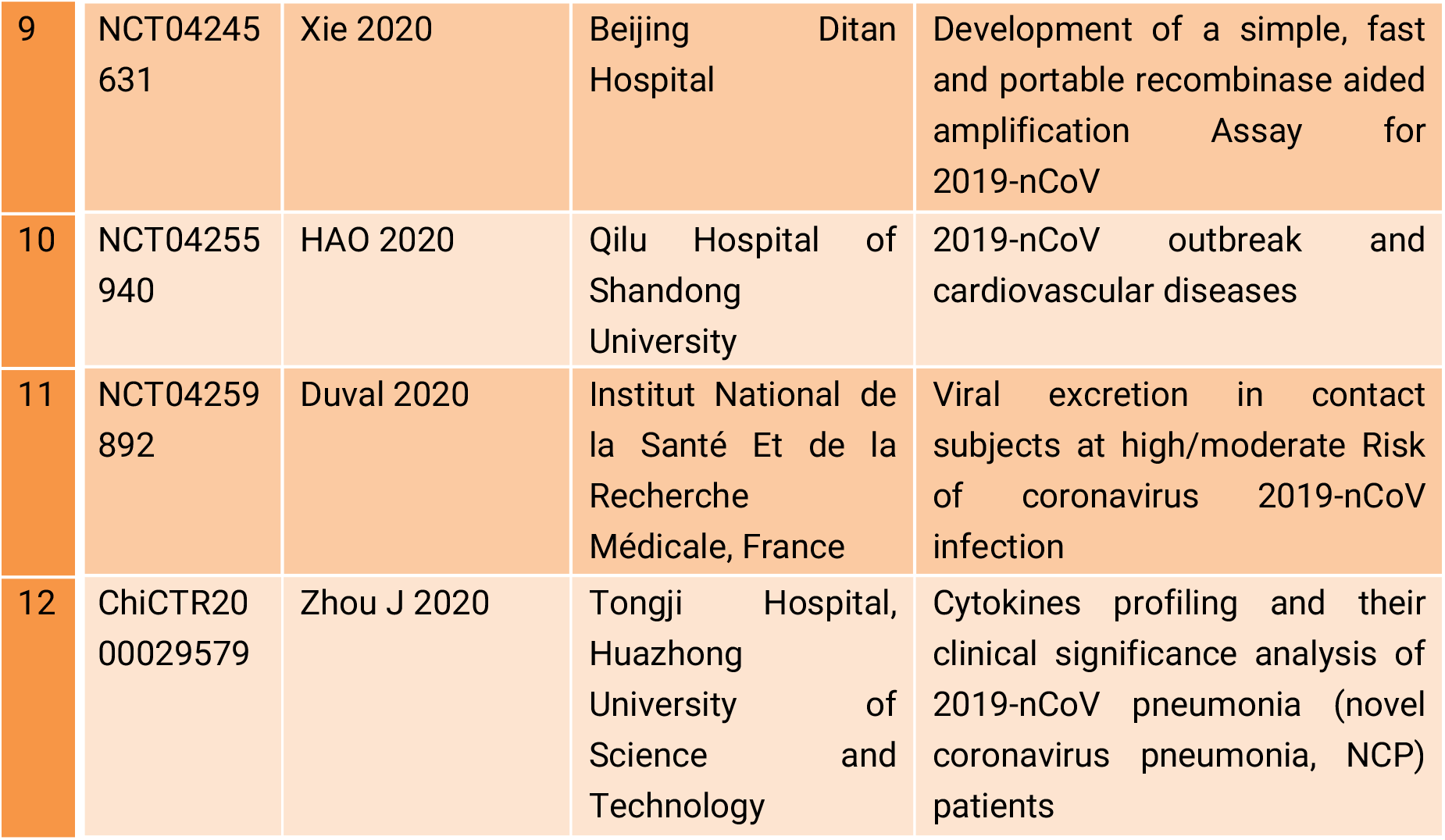
Summary of observational registered clinical trials.

| No | Register number | Study leader (year) | Primary sponsor | Study name |
| --- | --- | --- | --- | --- |
| 1 | ChiCTR2000029637 | Zhang Z 2020a | Guangdong Provincial Hospital of Chinese Medicine | An observational study for Xin-Guan-1 formula in the treatment of 2019-nCoV pneumonia (novel coronavirus pneumonia, NCP) |
| 2 | ChiCTR2000029430 | Zhang Z 2020b | Hubei Integrated Hospital of Traditional Chinese and Western Medicine | Study for the TCM syndrome characteristics of novel coronavirus pneumonia (COVID-19) |
| 3 | ChiCTR2000029462 | Li J 2020 | The First Affiliated Hospital of He'nan University of Chinese Medicine | Study for clinical characteristics and distribution of TCM syndrome of 2019-nCoV pneumonia (novel coronavirus pneumonia, NCP) |
| 4 | ChiCTR2000029437 | Xia W 2020 | Hubei Provincial Integrated Hospital of traditional Chinese and Western Medicine | A single arm study for combination of traditional Chinese and Western Medicine in the treatment of novel coronavirus pneumonia (COVID-19) |
| 5 | ChiCTR2000029592 | Zheng X 2020 | Union Hospital, Tongji Medical College, Huazhong University of Science and Technology | Study for Arbidol Hydrochloride in the Prophylaxis of Novel Coronavirus pneumonia in High-risk Population with History of Exposed to 2019-nCoV pneumonia |
| 6 | ChiCTR2000029624 | Lu H 2020 | Shanghai Public Health Clinical Center | A real world study for traditional Chinese Medicine in the treatment of 2019-nCoV pneumonia (novel coronavirus pneumonia, NCP) |
| 7 | NCT04262921 | Yazdan 2020 | Institut National de la Santé Et de la Recherche Médicale, France | Clinical Characterization Protocol for Severe Emerging Infections |
| 8 | NCT04256395 | Dong 2020 | Beijing Tsinghua Chang Gung Hospital | Efficacy of a self-test and self-alert mobile applet in detecting susceptible infection of 2019-nCoV |
| 9 | NCT04245631 | Xie 2020 | Beijing Ditan Hospital | Development of a simple, fast and portable recombinase aided amplification Assay for 2019-nCoV |
| 10 | NCT04255940 | HAO 2020 | Qilu Hospital of Shandong University | 2019-nCoV outbreak and cardiovascular diseases |
| 11 | NCT04259892 | Duval 2020 | Institut National de la Santé Et de la Recherche Médicale, France | Viral excretion in contact subjects at high/moderate Risk of coronavirus 2019-nCoV infection |
| 12 | ChiCTR2000029579 | Zhou J 2020 | Tongji Hospital, Huazhong University of Science and Technology | Cytokines profiling and their clinical significance analysis of 2019-nCoV pneumonia (novel coronavirus pneumonia, NCP) patients |

**Figure 1.**
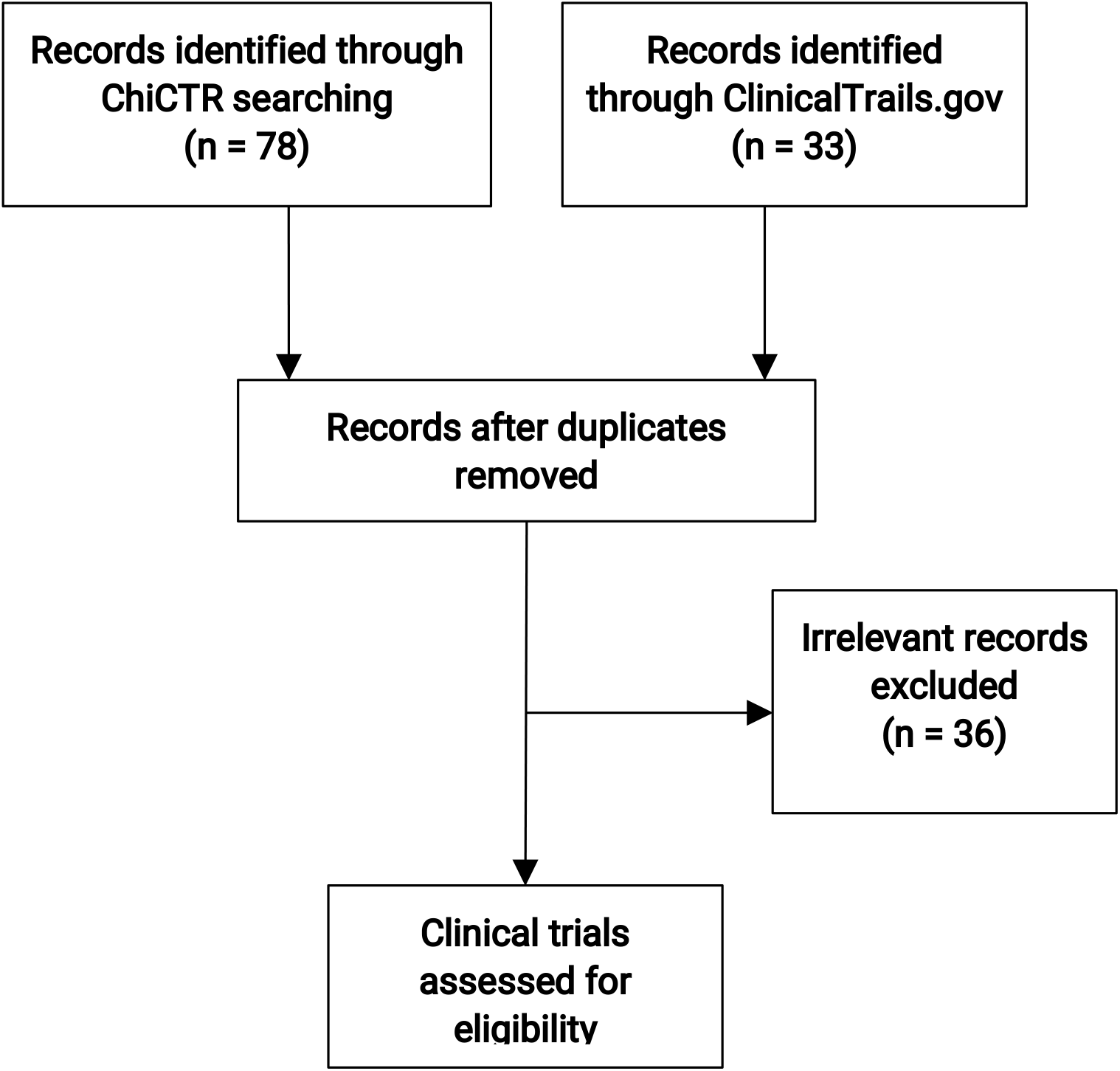
The flowchart of retrieval of the registered clinical trials.

### General characteristics of the clinical trials

The trials were sponsored by Chinese organizations, except for two from France (NCT04262921, NCT04259892). The following organizations sponsored more than three trials: Tongji Hospital, Tongji Medical College, Huazhong University of Science and Technology; The First Affiliated Hospital, College of Medicine, Zhejiang University; Xinhua Affiliated Hospital, Hubei University of Chinese Medicine; Zhejiang Chinese Medical University; Shanghai Public Health Clinical Center; and Hospital of Chengdu University of Traditional Chinese Medicine (Figure 2). The study sponsors belonged to different regions such as Hubei, Beijing, Zhejiang, Guangdong, Sichuan, Shanghai, etc. From the perspective of research type, most of them were interventional studies mainly aiming at drug therapy, and 12 were observation studies.

**Figure 2.**
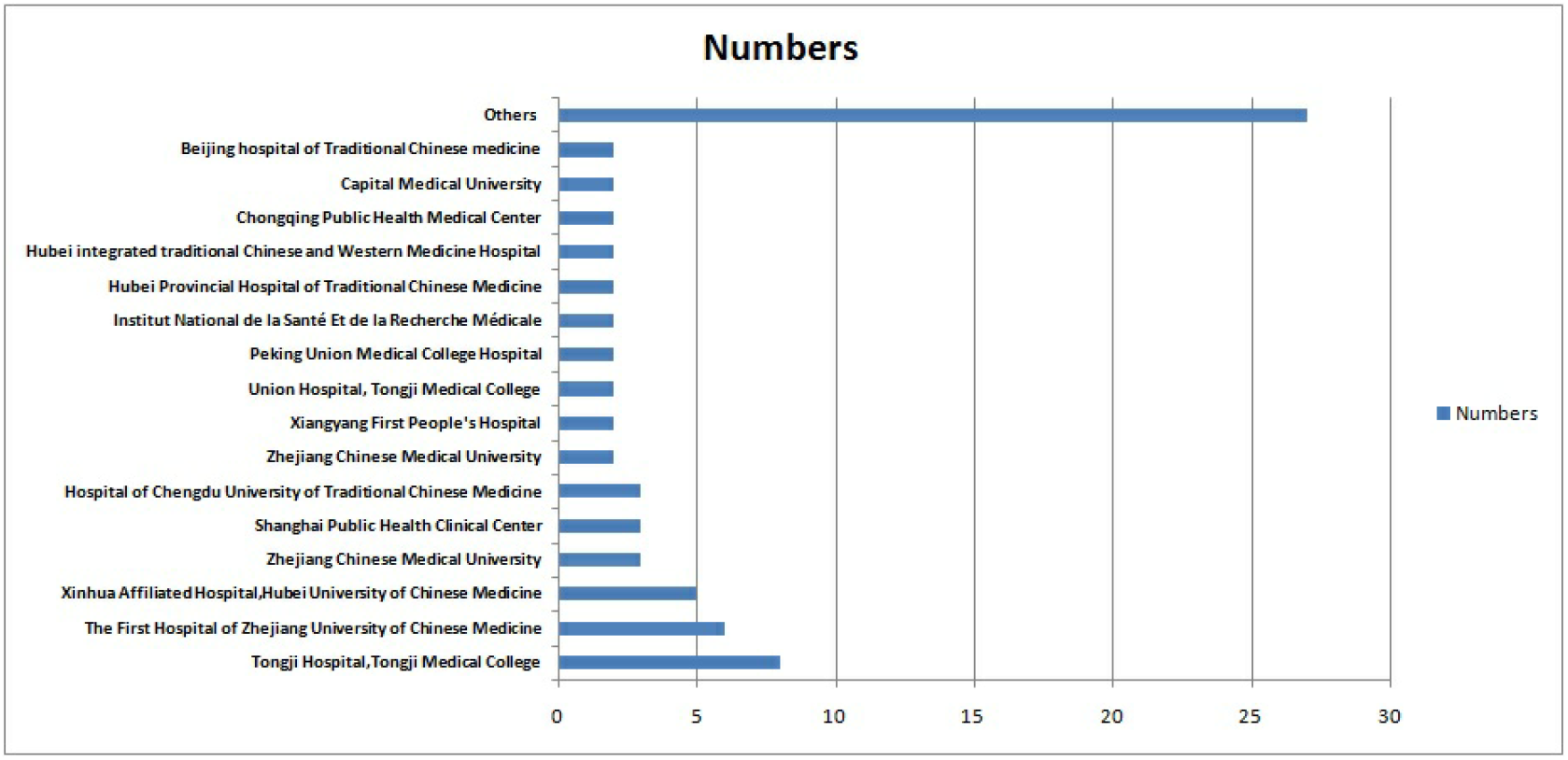
The primary sponsors of the registered clinical trials.

Most of the trials have passed the ethical review, whereas some are still in the preparation stage and only 11 trials have just started to recruit patients, however, none of the registered clinical trials have been completed. The first trial registered on January 23, 2020 was a randomized controlled trial of “A randomized, open-label, blank-controlled trial for the efficacy and safety of lopinavir-ritonavir and interferon-alpha 2b in hospitalization patients with novel coronavirus pneumonia (COVID-19)”, which was sponsored by the Wuhan Jinyintan Hospital.

In terms of trial stages, 34 trials were exploratory or in the preliminary experiment stage (phrase 0 clinical trial), 15 studies were in the extended validation stage with indications of drugs in the market (phrase IV), only 4 trials in phrase III (“NCT04252664, Mild/Moderate 2019-nCoV Remdesivir RCT” and NCT04257656, Severe 2019-nCoV Remdesivir RCT” by Cao B *et al*; “NCT04252274, Efficacy and Safety of Darunavir and Cobicistat for Treatment of Pneumonia Caused by 2019-nCoV and NCT04261517, Efficacy and Safety of Hydroxychloroquine for Treatment of Pneumonia Caused by 2019-nCoV (HC-nCoV) by Lu H *et al*; each individual study included hundreds of samples”. However, other studies belonged to the unspecified items.

The median sample size was 100 (IQR: 60 – 200), and the median execute time of the studies was 179 d (IQR: 94 – 366 d). General characteristics of the clinical trials were summarized in Table 3 and Table 4.

**Table 3.** Characteristics of the included interventional trials and participants.

| No | Register number | Study phrase | Start time | End time | Time consuming (days) | Sample | Intervention | Control |
| --- | --- | --- | --- | --- | --- | --- | --- | --- |
| 1 | ChiCTR2000029638 | 0 | 2020-02-03 | 2020-08-01 | 179 | 100 | Nebulization of recombinant super-compound interferon (rSIFN-co) | Nebulization of interferon α |
| 2 | ChiCTR2000029387 | N/A | 2020-01-25 | 2021-01-25 | 367 | 108 | Group A: Ribavirin + Interferon alpha-1b<br>Group B: lopinavir / ritonavir + interferon alpha-1b<br>Group C: Ribavirin + LPV/r+Interferon alpha-1b |  |
| 3 | ChiCTR2000029386 | N/A | 2020-01-29 | 2021-01-29 | 367 | 48 | Methylprednisolone, intravenous injection | Without any glucocorticoid therapy |
| 4 | ChiCTR2000029435 | 0 | 2020-02-01 | 2020-03-31 | 60 | 40 | TCM intervention | Placebo |
| 5 | ChiCTR2000029308 | N/A | 2020-01-10 | 2021-01-10 | 367 | 160 | Lopinavir-ritonavir tablets (each containing 200 mg of lopinavir and 50 mg of ritonavir) | Conventional standardized treatment |
| 6 | ChiCTR2000029400 | 0 | 2020-01-29 | 2020-12-31 | 338 | 60 | Group A: traditional Chinese medicine treatment<br>Group B: Lopinavir/Ritonavir<br>Group C: traditional Chinese medicine treatment + Lopinavir/Ritonavir |  |
| 7 | ChiCTR2000029418 | 0 | 2020-02-03 | 2020-08-31 | 211 | 42 | Combined Treatment of Chinese medicine and western medicine | Western medicine treatment |
| 8 | NCT04244591 | N/A | 2020-1-26 | 2020-4-25 | 91 | 80 | Methylprednisolone 40 mg q12h for 5 d | Standard care |
| 9 | NCT04251871 | N/A | 2020-1-22 | 2021-1-22 | 367 | 150 | Conventional medicines and Traditional Chinese Medicines (TCMs) granules | Conventional medicines |
| 10 | ChiCTR2000029436 | 0 | 2020-02-01 | 2020-12-31 | 335 | 100 | TCM syndrome differentiation treatment+ Western medicine treatment | Western medicine treatment |
| 11 | ChiCTR2000029432 | 0 | 2020-02-01 | 2020-04-30 | 90 | 72 | Tanreqing injection | No |
| 12 | ChiCTR2000029431 | 0 | 2020-01-29 | 2021-12-31 | 338 | 45 | Group A: Ankylosaurus<br>Group B: Ankylosaurus+M1 suppression therapy<br>Group C: Critical Treatment in Critical Period |  |
| 13 | ChiCTR2000029381 | 4 | 2020-01-01 | 2020-12-31 | 366 | 400 | XuebijingInjection | Conventional treatment |
| 14 | ChiCTR2000029487 | N/A | 2020-02-10 | 2020-03-31 | 51 | 200 | Isolation and oral GubiaoJiedu Ling Chinese medicine | Isolated observation |
| 15 | ChiCTR2000029479 | N/A | 2020-01-30 | 2020-05-01 | 93 | 10,000 | Jinhao Artemisia Antipyretic Granules | non-intervention |
|  |  |  |  |  |  |  | Huoxiangzhengqi |  |
| 16 | ChiCTR2000029468 | N/A | 2020-02-01 | 2020-06-30 | 151 | 120 | Lopinavir/litonavir (LPV/r)+emtricitabine (FTC)/Tenofovir alafenamide Fumarate tablets (TAF) in combination | Lopinavir / ritonavir |
| 17 | ChiCTR2000029461 | 0 | 2020-02-03 | 2021-12-31 | 698 | 100 | TCM decoctions+ basic conventional therapy | Basic conventional therapies |
| 18 | ChiCTR2000029460 | 0 | 2020-02-03 | 2021-12-31 | 698 | 100 | Shadowboxing +conventional treatment | Conventional treatments |
| 19 | ChiCTR2000029459 | 0 | 2020-02-03 | 2021-12-31 | 698 | 100 | Pulmonary rehabilitation+ Conventional treatment | Conventional treatment |
| 20 | ChiCTR2000029439 | 0 | 2020-02-01 | 2021-12-31 | 700 | 120 | TCM standard decoctions + basic western medical therapies | Basic western medical therapies |
| 21 | ChiCTR2000029438 | 4 | 2020-02-01 | 2021-12-01 | 670 | 100 | Conventional medicine + TCM | Western medical therapies |
| 22 | NCT04252274 | 3 | 2020-1-31 | 2020-12-31 | 336 | 30 | Darunavir, Cobicistat + conventional treatments | Darunavir and Cobicistat |
| 23 | NCT04261517 | 3 | 2020-2-6 | 2020-12-31 | 330 | 30 | Hydroxychloroquine and conventional treatments | Conventional treatments |
| 24 | ChiCTR2000029544 | 0 | 2020-02-04 | 2020-05-31 | 118 | 20 | Current antiviral treatment+ BaloxavirMarboxil tablets | Current antiviral treatment |
| 25 | ChiCTR2000029542 | 4 | 2020-02-03 | 2020-07-30 | 179 | 20 | Chloroquine | Conventional management |
| 26 | ChiCTR2000029541 | N/A | 2020-02-01 | 2020-12-01 | 305 | 80 | DRV/c + Conventional treatment containing thymosin | LPV/r + Conventional treatment containing thymosin |
| 27 | ChiCTR2000029539 | 0 | 2020-02-03 | 2021-02-02 | 366 | 328 | Conventional standardized treatment and Lopinavir-Ritonavir | Conventional standardized treatment |
| 28 | ChiCTR2000029518 | 0 | 2020-02-04 | 2020-04-30 | 87 | 80 | Ordinary Chinese and Western Medicine | Ordinary Western medicine |
| 29 | ChiCTR2000029517 | 0 | 2020-02-04 | 2020-04-30 | 87 | 100 | Chinese medicine decoction | Placebo |
| 30 | ChiCTR2000029496 | 4 | 2020-01-29 | 2021-01-29 | 367 | 60 | Routine medical treatment+ Novaferon Atomization inhalation | On the basis of routine medical treatment, the patients were given lopinavir/ritonavir tablets (Kaletra) |
| 31 | ChiCTR2000029495 | 0 | 2020-02-03 | 2021-12-31 | 698 | 60 | Traditional Chinese Medicine+ psychological intervention | Traditional Chinese Medicine |
| 32 | ChiCTR<br>200002<br>9493 | 0 | 2020-02<br>-03 | 2021-12<br>-31 | 698 | 100 | TCM<br>decoctions+<br>basic<br>western<br>medical<br>therapies | Basic western<br>medical<br>therapies |
| 33 | ChiCTR<br>200002<br>9580 | 0 | 2020-01<br>-31 | 2020-12<br>-31 | 335 | 70 | Ruxolitinib<br>combined<br>with<br>mesenchym<br>al stem cell | Routine<br>treatment |
| 34 | ChiCTR<br>200002<br>9589 | 0 | 2020-02<br>-05 | 2021-12<br>-31 | 330 | 60 | Reduning<br>injection<br>combined<br>with basic<br>western<br>medical<br>therapies | Basic western<br>medical<br>therapies |
| 35 | ChiCTR<br>200002<br>9600 | 0 | 2020-01<br>-30 | 2020-04<br>-29 | 91 | 60 | Lopinavir<br>and<br>Ritonavir +<br>alpha-Interfe<br>ron<br>atomization | Alpha-Interferon<br>atomization |
| 36 | ChiCTR<br>200002<br>9601 | 0 | 2020-02<br>-01 | 2020-08<br>-01 | 183 | 400 | Health<br>education+<br>Basic<br>treatment of<br>Western<br>medicine+<br>Dialectical<br>treatment of<br>traditional<br>Chinese<br>medicine | Health<br>education+<br>Basic treatment<br>of western<br>medicine |
| 37 | ChiCTR<br>200002<br>9602 | 0 | 2020-02<br>-01 | 2020-08<br>-01 | 183 | 600 | Health<br>education,<br>follow-up<br>condition<br>managemen<br>t by team of<br>family<br>doctors +<br>Chinese | Health<br>education,<br>follow-up<br>condition<br>management<br>by team of<br>family<br>doctors |

|  |  |  |  |  |  |  | medicine<br>treatment |  |
| --- | --- | --- | --- | --- | --- | --- | --- | --- |
| 38 | ChiCTR<br>200002<br>9603 | 0 | 2020-02<br>-06 | 2020-05<br>-31 | 116 | 160 | Conventional<br>standardized<br>treatment<br>and<br>ASC09/Ritonavir | Conventional<br>standardized<br>treatment +<br>Lopinavir/Ritonavir |
| 39 | ChiCTR<br>200002<br>9605 | 4 | 2020-02<br>-05 | 2021-02<br>-05 | 367 | 200 | Low dose ,<br>Medium<br>dose and<br>High dose<br>of<br>Shuanghuan<br>glian | Routine<br>treatment |
| 40 | ChiCTR<br>200002<br>9578 | 0 | 2020-02<br>-06 | 2020-04<br>-30 | 85 | 10,000 | Integrated<br>Traditional<br>Chinese and<br>Western<br>Medicine | No |
| 41 | ChiCTR<br>200002<br>9573 | 4 | 2020-02<br>-05 | 2020-06<br>-30 | 147 | 200 | Novaferon<br>injection +<br>atomized<br>inhalation +<br>Arbidol<br>Tablets | Arbidol Tablets |
| 42 | ChiCTR<br>200002<br>9572 | 0 | 2020-02<br>-05 | 2021-04<br>-30 | 86 | 30 | Conventional<br>treatment<br>combined<br>with<br>umbilical<br>cord blood<br>mononuclear<br>cells group | Conventional<br>treatment |
| 43 | ChiCTR<br>200002<br>9569 | 0 | 2020-02<br>-05 | 2021-04<br>-30 | 86 | 30 | Conventional<br>treatment<br>combined<br>with<br>umbilical<br>cord<br>mesenchymal<br>stem cell<br>conditioned medium group | Conventional<br>treatment |
| 44 | ChiCTR<br>200002<br>9559 | 4 | 2020-01<br>-31 | 2020-02<br>-29 | 30 | 200 | Group 1:<br>Hydroxychloroquine 0.1<br>oral 2/d<br>Group 2:<br>Hydroxychloroquine 0.2<br>oral 2/d | Starch pill oral<br>2/ day |
| 45 | ChiCTR<br>200002<br>9558 | 0 | 2020-01<br>-29 | 2020-05<br>-01 | 94 | 200 | Chinese<br>medicine<br>treatment<br>combined<br>with<br>Western<br>medicine<br>treatment | No |
| 46 | ChiCTR<br>200002<br>9550 | N/A | 2020-01<br>-29 | 2020-05<br>-01 | 94 | 300 | Compound<br>Yinchai<br>granules<br>15g,<br>Qingqiao<br>antiviral<br>granules<br>15g, tid, with<br>warm water | Compound<br>Yinchai granules<br>15g, Qingqiao<br>antiviral<br>granules 15g,<br>q4hwith warm<br>water |
| 47 | ChiCTR<br>200002<br>9549 | N/A | 2020-02<br>-03 | 2020-05<br>-01 | 89 | 200 | Western<br>medicine<br>routine<br>treatment<br>combined<br>with<br>traditional<br>Chinese<br>medicine<br>treatment | Western<br>medicine routine<br>treatment |
| 48 | ChiCTR<br>200002<br>9548 | 0 | 2020-02<br>-04 | 2020-06<br>-03 | 121 | 30 | Group A: BaloxavirMarboxil<br>Group B: Favipiravir<br>Group C: Lopinavir-Ritonavir |  |
| 49 | NCT042<br>60594 | 4 | 2020-02<br>-07 | 2020-12<br>-30 | 326 | 380 | Arbidol<br>tablets +<br>basic<br>treatment | Basic treatment |
| 50 | NCT04261907 | N/A | 2020-02-07 | 2020-06-30 | 145 | 160 | ASC09/ritonavir + conventional standardized treatment | Lopinavir/ritonavir tablet+ conventional standardized treatment |
| 51 | NCT04257656 | 3 | 2020-02-06 | 2020-04-03 | 58 | 453 | active remdesivir | Placebos matched remdesivir |
| 52 | ChiCTR2000029636 | 0 | 2020-02-07 | 2020-07-30 | 175 | 40 | Conventional standardized treatment+ vMIP atomized inhalation | No |
| 53 | ChiCTR2000029626 | 0 | 2020-02-17 | 2020-08-01 | 167 | 20 | N/A | N/A |
| 54 | ChiCTR2000029621 | 4 | 2020-01-01 | 2020-12-31 | 365 | 380 | Arbidol tablets + basic treatment | Basic treatment |
| 55 | ChiCTR2000029609 | 4 | 2020-02-10 | 2020-12-31 | 324 | 177 | Group 1: mild-moderate chloroquine<br>Group 2: mild-moderate Lopinavir/ritonavir<br>Group 3: mild-moderate combination<br>Group 4: severe-chloroquine<br>Group 5: severe- Lopinavir/ritonavir |  |
| 56 | ChiCTR2000029606 | 0 | 2020-01-15 | 2022-12-31 | 880 | 63 | Group A: Conventional treatment followed by Intravenous infusion of Human Menstrual Blood-derived Stem Cells preparations<br>Group A control: Conventional treatment<br>Group B1: Artificial liver therapy+ conventional treatment |  |
|  |  |  |  |  |  |  | Group B2: Artificial liver therapy followed by Intravenous infusion of Human Menstrual Blood-derived Stem Cells preparations+ conventional treatment<br>Group B control: Conventional treatment |  |
| 57 | ChiCTR 200002 9625 | 0 | 2020-02-17 | 2020-08-01 | 167 | 80 | inapplicable | inapplicable |
| 58 | NCT042 63402 | 4 | 2020-02-01 | 2020-07-01 | 152 | 100 | Basic symptomatic supportive treatment +methylprednisolone (<40mg/d intravenous drip for 7 ) | Basic symptomatic supportive treatment +methylprednisolone (40-80 mg/d intravenous drip for 7 d) |
| 59 | NCT042 54874 | 4 | 2020-02-01 | 2020-07-01 | 152 | 100 | Abidol Hydrochloride combined with interferon atomization | Abidol hydrochloride |
| 60 | NCT042 61270 | N/A | 2020-02-01 | 2020-07-01 | 152 | 180 | Group 1: ASC09F + Oseltamivir<br>Group 2: Ritonavir+ Oseltamivir<br>Group 3: Oseltamivir |  |
| 61 | NCT042 52664 | 3 | 2020-02-01 | 2020-04-07 | 75 | 308 | Remdesivir | Remdesivir placebo |
| 62 | NCT042 61426 | 2,3 | 2020-2-10 | 2020-06-30 | 142 | 80 | IVIG therapy+ standard care | Standard care |
| 63 | NCT042 55017 | 4 | 2020-02-01 | 2020-07-01 | 152 | 400 | Group 1: Abidol hydrochloride<br>Group 2: Oseltamivir | Symptomatic supportive treatment |
|  |  |  |  |  |  |  | Group 3:<br>Lopinavir/ritonavir |  |

**Table 3.** The methodology quality of the observational trials using Newcastle-Ottawa scale.

| Register number | Representativeness of the exposed cohort | Select ion of the non exposed cohort | Ascertain ment of exposure | Demonstr ation that outcome of interest was not present at start of study | Comparability of cohorts on the basis of the design or analysis | Assessment of outcome | Was follow-up long enough for outcomes to occur | Adequacy of follow up of cohort s | Score s |
| --- | --- | --- | --- | --- | --- | --- | --- | --- | --- |
| ChiCTR2000029637 | 1 | 1 | 1 | 1 | 1 | 0 | 1 | 0 | 6 |
| ChiCTR2000029430 | 1 | 1 | 1 | 1 | 1 | 0 | 0 | 0 | 5 |
| ChiCTR2000029462 | 1 | 1 | 1 | 1 | 1 | 1 | 0 | 0 | 6 |
| ChiCTR2000029437 | 1 | 1 | 1 | 1 | 1 | 1 | 0 | 0 | 6 |
| ChiCTR2000029592 | 1 | 1 | 1 | 1 | 1 | 1 | 0 | 0 | 6 |
| ChiCTR2000029624 | 1 | 1 | 1 | 1 | 1 | 1 | 0 | 0 | 6 |
| NCT04262921 | 1 | 1 | 1 | 1 | 1 | 0 | 1 | 0 | 6 |
| NCT04256395 | 1 | 1 | 1 | 1 | 1 | 0 | 1 | 0 | 6 |
| NCT04245631 | 1 | 1 | 1 | 1 | 0 | 0 | 1 | 0 | 5 |
| NCT04255940 | 1 | 1 | 1 | 1 | 0 | 0 | 0 | 0 | 4 |
| NCT04259892 | 1 | 1 | 1 | 1 | 1 | 0 | 1 | 0 | 6 |
| ChiCTR2000029579 | 1 | 1 | 1 | 1 | 1 | 0 | 0 | 0 | 5 |

Note: A study can be awarded a maximum of one point for each numbered item within the Selection and Outcome categories. A maximum of two points can be given for Comparability.

**Table 4.**
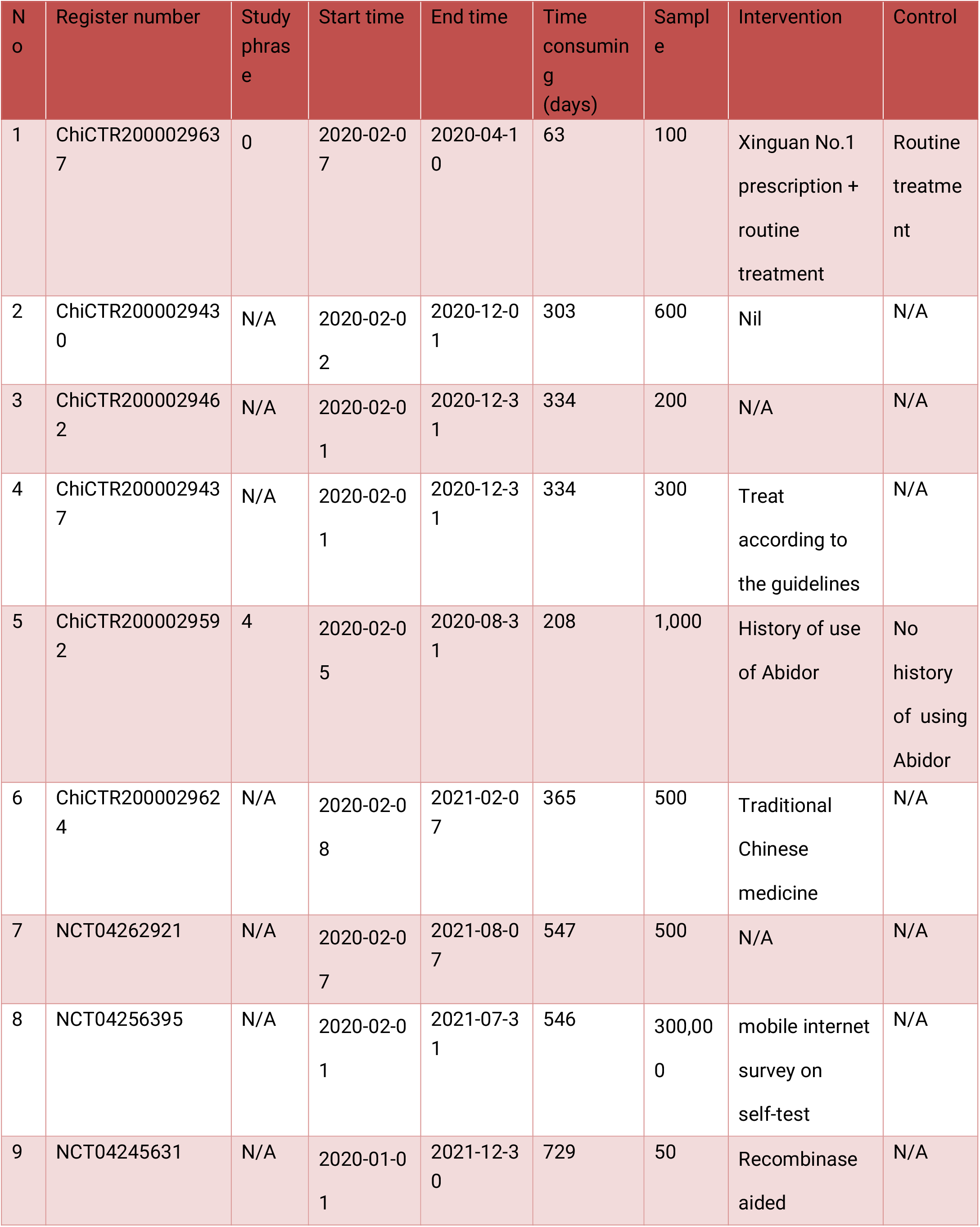
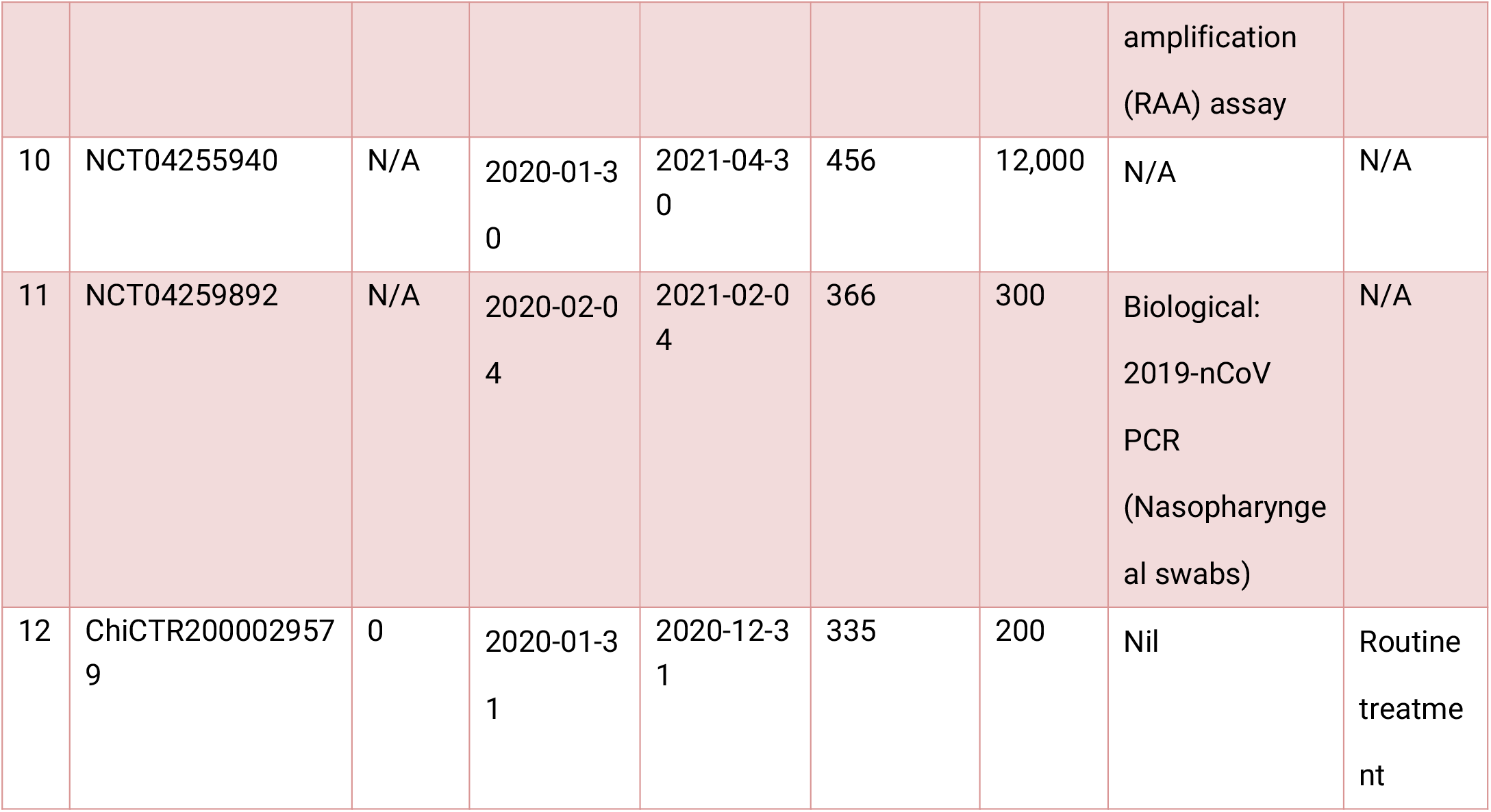
Summary of observational registered clinical trials.

### Characteristics of inclusion criteria

The common characteristics of inclusion criteria included: signing informed consent; age over 18 years; laboratory test (RT-PCR) confirmed infection of COVID-19 (diagnostic criteria for pneumonia diagnosis in line with “Protocol of Prevention and Control of Novel Coronavirus Pneumonia”); chest imaging confirmed lung involvement; participants were willing to be assigned to any designated treatment group randomly; and participants agreed not to participate in another study of the investigator until completion of the present study Most of the studies were limited to light subjects (ordinary subjects), and few of the studies included severe patients.

### Characteristics of exclusion criteria

The common characteristics of the exclusion criteria were: critical patients with COVID-19; pregnant and lactating women; allergic patients; patients with tumors or serious heart, brain, kidney, and hemoglobin disease and other diseases; patients with mental disorders, drug abuse or dependence history; subjects who failed to get informed consent; and researchers’ opinion that the subject is not suitable for the study.

### Intervention and comparison

The main intervention methods of registered clinical trials included treatment with traditional Chinese medicine, western medicine, and integrated traditional Chinese and western medicine. The outcomes of treatment observation mainly included clinical rehabilitation time, incidence of using mechanical ventilation, incidence in ICU, mortality, all kinds of complications and virological detection indicators, etc. The medication methods mainly included oral, injection, atomization inhalation, etc.; the medication time was generally more than one week. The time period of outcome was more than 2–4 weeks. The controls were treated either with placebo or routine treatment.

Among the registered clinical trials, 30 were Western medicine-based treatments, and the methods of intervention mainly included: i) antiviral drugs, such asrhetcivir, Abidol, fabiravir, chloroquine phosphate, asc09/ritonavir compound tablets, lopinavir/ritonavir (Coriolus), hydroxychloroquine, chloroquine, baloxavir, darunavir/Corbis, etutabine/propofol tenofovir, etc.; ii) antiviral drugsin combination with biological agents, such aslucotinib combined with mesenchymal stem cell therapy, recombinant cytokine gene derived protein injection combined with Abidol or lopinavir/ritonavir, recombinant virus macrophage inflammatory protein for aerosol inhalation injection or lopinavir/ritonavir tablets combined with thymosin A1, and lopinavir/ritonavir combined with interferon-α2b; iii) biological agents (products), such as uterine blood stem cells, interferon, cord blood mononuclear cells, cord mesenchymal stem cell conditioned medium, recombinant cytokine gene-derived protein, immunoglobulin, etc.; and iv)steroid therapy, for example, glucocorticoids (intervention in critical patients).

There were 26 registered clinical trials with traditional Chinese medicine treatment. Traditional Chinese medicine treatment drugs were mainly various kinds of Chinese herbal medicines (decoction, capsule, granule, etc.), including Feiyanyihao, Qingfeijiedutang, Xinguanyihao, Lianhuaqingwen capsule, etc. The main ingredients of these drugs included antiviral and immunomodulatory Chinese herbal formulas. In addition, traditional Chinese medicine treatment also involved certain traditional Chinese medicine injection, such as Xuebijin Injection, Shuanghuanglian injection and Tanreqing injection.

There were 19 registered clinical trials using a combination treatment of Chinese and western medicine, and the intervention included the combination of the above mentioned Chinese herbs and western antiviral drugs.

### Outcomes and timing of measurement

The outcomes mainly included: clinical symptoms, mortality, chest CT, viral nucleic acid detection, body temperature, clinical improvement, critically ill patients (%); lung function; the time to 2019-nCoV RNA negativity in patients, time for lung recovery, mechanical ventilation time; length of stay in hospital, time for body temperature recovery, inflammatory cytokines, SOFA score, St Georges respiratory questionnaire, SGRQ, modified Barthel Index, MBI, and incidence of adverse events. Additionally, some other laboratory tests for novel coronavirus were also selected, including routine blood test, routine urine test, C-reactive protein, procalcitonin, erythrocyte sedimentation rate, muscle enzyme, troponin, myoglobin, D dimer, blood gas analysis, coagulation routine, new coronavirus nucleic acid test, T cell subgroup analysis, hospitalization period etc.

The follow-up period of the outcome measure was mostly 2–4 weeks, but some studies did not set forth a plan.

### Methodology quality

According to the Cochrane bias risk assessment results (Figure 3), the quality assessment of the interventional study methodology is generally low. Most trials reported randomization, while the other trials had high risk of biases in randomization (17 trials did not mention randomization and 6 trials were judged as non-randomized trials); Few trials conducted distribution concealment; only nine trials implemented blinding of participants, personnel and outcome assessment; None of the 63 trials clarified drop-out and follow-up bias. However, other bias risks, such as the risk of conflict of interest among drug manufacturers, are unclear.

**Figure 3.**
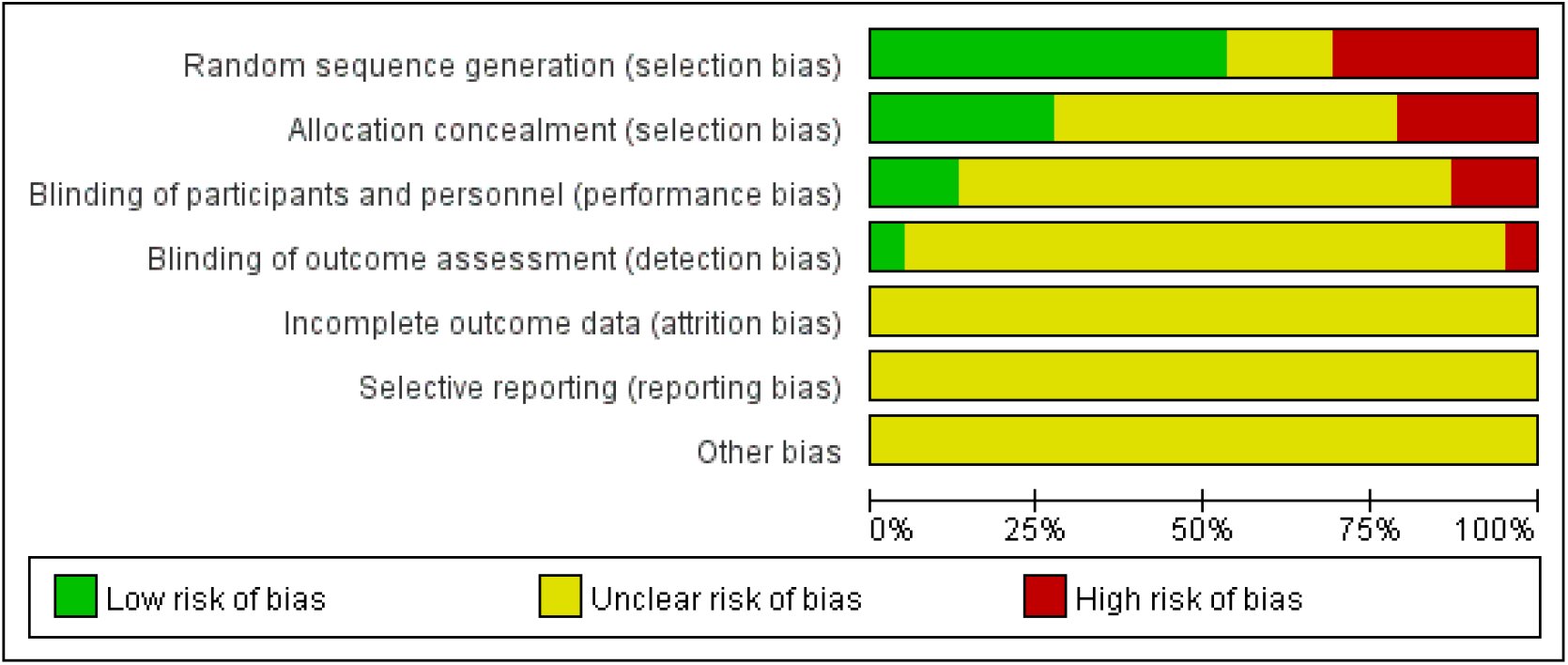
Risk of bias graph across all included interventional clinical trials.

The NOS scores of the observational trials are from 4 to 6 (Table 3). Most of the observational trials have high risk of biases in assessment outcome, follow-up of outcome and adequacy of follow up of cohorts (Figure 4). Therefore, the overall quality of registered observational trials is low.

**Figure 4.**
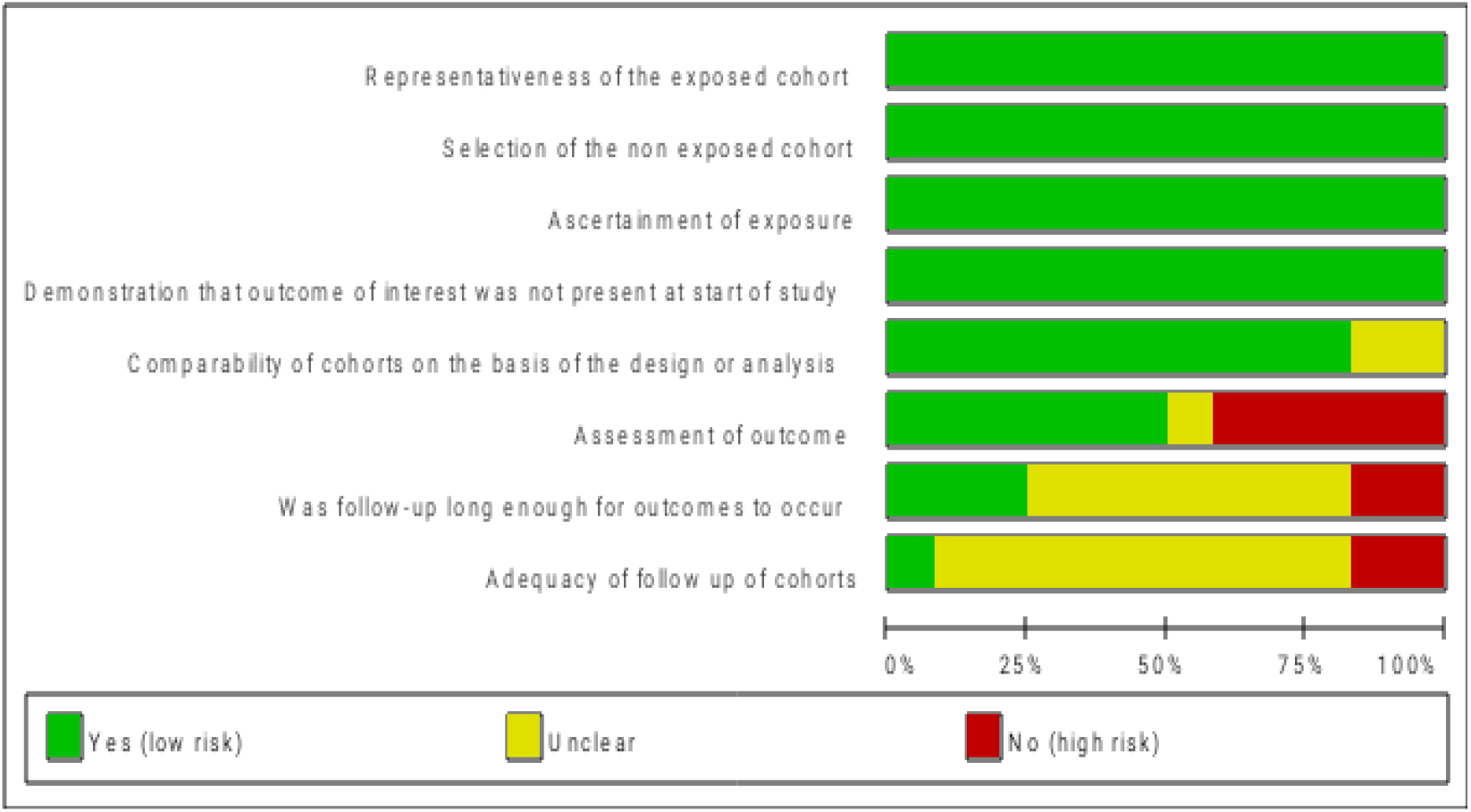
Risk of bias graph across all included observational studies.

## Discussion

COVID-19, being a new and poorly understood infectious disease, has no recognized effective treatment strategy. Coronavirus outbreak has caused great harm to China and seriously threatened people’s health [21-23]. To deal with the disease, many intensive clinical trials have been carried out. Database search results indicated that current studies were mainly from China, involving the treatment with traditional Chinese medicine, Western medicine, and the combination of traditional Chinese and Western medicine and the primary sponsors were mainly the hospitals of China. However, the median sample size of the trials was 100 (IQR: 60 – 200) and most trials had a small sample size. Hence, the future evidence level of these studies is low.

According to the summary results, only 11 trials have begun to recruit patients, and none of the registered clinical trials had been completed. Of them, 34 trials were early clinical exploratory trials or in a pre-experiment stage (phrase 0). Fifteen trials belonged to phrase IV and some drugs that have been licensed for other diseases such as chloroquine phosphate, abidol, fabiravir, asc09/ritonavir compound tablets, lopinavir/ritonavir, hydroxychloroquine, chloroquine, etc. were used in registered clinical trials of COVID-19. Four studies were phase III clinical trials with remdesivir, darunavir and cobicistat, and hydroxychloroquine.

The main methods of intervention included traditional Chinese medicine involving 26 trials, Western medicine involving 30 trials, and integrated traditional Chinese medicine and Western medicine involving 19 trials.

At present, conventional treatment strategies for COVID-19 mainly involve use of antivirals, improving patients’ immunity, intervening autoimmune damage (against immune storm caused by cytokines), and symptomatic treatment. Western drugs have been shown to be superior to traditional Chinese medicine in terms of *in vitro* antiviral effect. However, as Chinese herbs have both antiviral and immunomodulatory effects based on low quality clinical evidence, they have the potential value in the prevention and treatment of COVID-19.

There were 26 registered clinical trials with traditional Chinese medicine treatment and 19 registered clinical trials using a combination treatment of Chinese and western medicine, suggesting that traditional Chinese medicine is a popular candidate for therapeutic drugs against COVID-19. At present, the combination of Chinese and western medicines (Qingfeipaidutang and chloroquine phosphate, abidol, lopinavir/ritonavir) is considered a better treatment strategy by experts, and has been listed in the “Protocol of Prevention and Control of Novel Coronavirus Pneumonia”; however, there is still a lack of high-quality evidence, and clinical verification is required.

Existing preliminary evidences suggest that the antiviral drug remdesivir (phase III clinical trials for light, moderate, and severe patients, expected to end on April 27, 2020) has a promising application prospect. The reasons are as follows: i) *in vitro* and in vivo cell test results indicated that even very low concentration of the drug has an antiviral effect [24, 25]; ii) animal trials have proved the drug safe for use [26]; and iii) “clinical tests indicate that the drug is effective against Ebola virus [27, 28]; iv) clinical case report is effective [29]. In addition, some of the validation drugs, such as chloroquine phosphate, Abidol, darunavir, and lopinavir/ritonavir (Coriolus Versicolor), have been proven safe and have shown strong antiviral potential *in vitro* [25]. Thus, these Western antiviral drugs have an application potential which needs to be verified in clinical practice.

In this review, we found that many trials used biological agents for immunotherapy of the disease. In light of the experience and lessons of severe acute respiratory syndrome (SARS) [30, 31], steroid therapy has been used cautiously in the treatment of COVID-19; therefore, we found only a few studies on steroid therapy.

From the perspective of inclusion and exclusion criteria, some people were excluded, such as children and adolescents, pregnant women, and patients with serious liver and kidney damage. Therefore, there is a lack of clinical evidence in this portion of the population.

The outcomes of clinical trial observation included clinical observation outcomes, physical examination and laboratory test results, however, some outcomes were subjective leading to measurement bias.

Based on Cochrane risk of bias items and NOS, we evaluated the quality of interventional trials and observational trials, respectively. The evaluation results showed that the overall quality of the registered clinical trials was low, indicating that most of the registered clinical studies had a greater risk of bias, and the level of evidence is relatively low in the future, which belittles the practice significance of the research. We believed that it is difficult to obtain reliable and high-quality evidence in near future. The main reasons for the low quality of the registered clinical trial protocols could be: i) insufficient clinical research ability of the researchers; and ii) researchers’ lack of experience in dealing with sudden health events.

We believed that it is necessary to improve the quality of research and to the registered clinical research programmes in strict accordance with the guidelines for clinical trials [32-35]. In addition, current clinical trials by different hospitals conducted spontaneously are not effectively organized and coordinated, so more scattered and disorderly. Some drugs that have not been tested *in vitro* or whose safety is of great concern are also being tested in clinical trials, which not only increase the risk of clinical trials, but also waste research resources. Hence, the administration of scientific research should strengthen their management and coordination and few promising drugs should be prioritized for clinical trials.

From these registered clinical studies, we found a serious limitation: most of the registered clinical research did not consider the “timeliness”, and still followed the conservative traditional study design paradigm. The median execute time (days) of the studies was 179 d (IQR: 94 – 366 d), which is highly unfavorable in the current critical situation. We believe that, in the current situation, the “timeliness” factor should be given importance in the design of clinical trials, so that the research does not lose its social significance. Therefore, in this critical situation, it is better to refer to the “sequential design” for clinical trials; “sequential design” not only requires small sample size, but also significantly shortens the research during, therefore, it is very conducive to the screening and discovery of drugs with significant efficacy [36, 37]. In addition, a very difficult problem is the treatment of severe and critical patients with COVID-19. For these patients, we suggested that: based on the “compassionate use drug” principle, with safe and obvious antiviral potential drugs, to conduct a staged small batch and single-arm clinical trials is feasible. We believed that “compassionate use drug” can not only meet the special needs of patients but also perform clinical effectiveness observation, research and analysis, so as to enhance the efficiency of research and benefit the patients [38-42]. Also, given a large number of clinical cases have accumulated information, and using available existing data for statistics and analysis with the help of new statistical methods such as clinical data-mining [43-45] and real-world study [46-48], etc., can help in quickly obtaining some very valuable information and save research time.

In brief, under the condition that there are a large number of cases to be selected at present, it is of great value for the treatment and prevention of COVID-19 to try to complete various clinical trial designs and data analysis scientifically and efficiently with a variety of clinical research designs and statistical analysis methods, and researchers should try in future.

## Conclusions

Disorderly and intensive clinical trials of COVID-19 using traditional Chinese medicine and Western medicine are ongoing or will be carried out in China. However, based on the poor quality and small sample size and long study execute period, we will not be able to obtain reliable, high-quality clinical evidence about COVID-19 treatment for quite a long time in the future. In order to effectively deal with the current sudden health emergencies, the National Administration of scientific research should strengthen their management and coordination to improve the study quality based on the guidelines for clinical trials. Also, it is important to ensure that some promising projects are prioritized.

In addition, we suggest that using a variety of study designs and statistical methods to scientifically and efficiently conduct the clinical trials, which has an extremely important value for the control of COVID-19.

## Data Availability

The data used to support the findings of this study are included within the study.

